# Early Insights from the Pumping Marvellous Home-Based Digital Cardiac Rehabilitation for Heart Failure in the UK

**DOI:** 10.1101/2025.09.11.25334847

**Authors:** Rajiv Sankaranarayanan, Nick Hartshorne-Evans, Kate Hornby, Matthew Sunter, Yvonne Millerick, Carys Barton, Ahmet Fuat, Dr Duwarakan Satchithananda, Fozia Ahmed, Patrick Doherty

## Abstract

**Background:** Cardiac rehabilitation (CR) uptake for heart failure (HF) in the UK remains low at around 15%, with evidence of disparities based on sex, ethnicity, socioeconomic status, availability of staffing and financial constraints. We analysed data from the Pumping Marvellous Foundation online home CR programme to understand who accesses this service and whether inequities persist, to help guide strategies to achieve equitable rehabilitation delivery.

**Methods:** The PMF online home CR platform was launched in August 2024 along with an educational booklet available for order (at no cost) by HF or CR teams. We analysed anonymised data (n=673) of registrants from August 2024 to July 2025. Variables analysed included demographics, ethnicity, deprivation index, HF type based on ejection fraction, referral source, time since diagnosis and prior CR participation. We also analysed CR booklet orders and assessed correlation with HF admissions as per national HF audit data. Descriptive statistics summarised distributions.

**Results:** 673 participants (median age 62 years; IQR 18 to 90 years, 12% aged >76 years) registered for the online CR classes from August 2024 to July 2025. The majority (63%) were women, and 6% were from minority ethnic communities. 35% of registrants had either mildly reduced (HFmrEF) or preserved (HFpEF) ejection fraction. 30% of referrals were from HF or CR teams, 29% obtained information via social media, and around 25% obtained information directly from PMF groups. There were also direct referrals from GPs (4%) and around 10% obtained referral information via Google search or YouTube. Analysis of the time since HF diagnosis demonstrated late entry to CR: 343 (51%) registered >12 months post-diagnosis, 88 (13%) within 3 months, and 130 (19%) within 6 to 12 months. Only 38 (6%) reported any prior CR participation. We also correlated CR booklet orders from hospitals with National HF Audit HF admissions. Within the limitations of the spread of the scatter, there was a general positive relationship: hospitals with more HF admissions tended to order more booklets. 33% of registrants came from the top 20 most deprived cities in England.

**Conclusions:** By providing free lifetime access to online cardiac rehabilitation, widening the access of cardiac rehab to more women and people without access to standard cardiac rehabilitation (due to staffing, cost constraints, accessibility issues) and reaching areas with socio-economic deprivation, the PMF online cardiac rehab platform can help to increase CR uptake and reduce the inequity in access to CR in the UK.

## Introduction

Heart failure (HF) affects approximately one million people in the UK, causing significant morbidity, mortality and healthcare costs [1] and the burden of HF is likely to increase [2]. Cardiac rehabilitation (CR) is a safe, multi-faceted intervention broadly encompassing exercise, patient education and support. It is recommended in national and international guidelines [3,4] and improves outcomes, including reduced hospitalisation, mortality and improved quality of life [5]. Yet, CR uptake for HF in the UK remains low at around 15% [5], with evidence of disparities based on sex, ethnicity, socioeconomic status, availability of staffing and financial constraints [5,6].

Digital and home-based CR programmes are emerging as scalable alternatives to hospital-based models, with potential to improve uptake and accessibility [3,7,8]. The Pumping Marvellous Foundation (PMF), a patient-led charity, developed an online home CR programme (“low-function” and “medium-function courses”) specifically for HF patients. This has been devised with expert input from HF specialists, cardiac rehab specialists, and HF patient experts. The portal also assessed registrants’ HF awareness levels through a quiz before and after completion of the program. The service was developed as an adjunct to existing CR services or in areas where CR is not commissioned for HF. We conducted this analysis to understand who accesses this service and whether inequities persist, to help guide strategies to achieve equitable rehabilitation delivery.

## Methods

The PMF online home cardiac rehabilitation platform was launched in August 2024 along with an educational booklet available for order (at no cost) by HF or CR teams. We analysed anonymised data (n=673) of registrants from August 2024 to July 2025. Variables analysed included demographics, ethnicity, deprivation index based on postcode and city, HF type based on ejection fraction, referral source, time since diagnosis, prior CR participation, and knowledge quiz scores (six-item multiple choice). We also analysed CR booklet orders and assessed correlation with HF admissions as per national HF audit data [9]. Descriptive statistics summarised distributions.

## Results

673 participants (median age 62 years; IQR 18-90 years, 12% aged >76 years) registered for the online cardiac rehabilitation classes from August 2024 to July 2025. The majority (63%) were women, and 6% were from minority ethnic communities. 35% of registrants had either mildly reduced (HFmrEF) or preserved (HFpEF) ejection fraction. Figure 1 illustrates the referral sources. ∼30% of referrals were from HF or CR teams, 29% obtained information via social media, and ∼25% obtained information directly from PMF groups. There were also direct referrals from GPs (4%) and around 10% obtained referral information via Google search or YouTube.

**Fig. 1.**
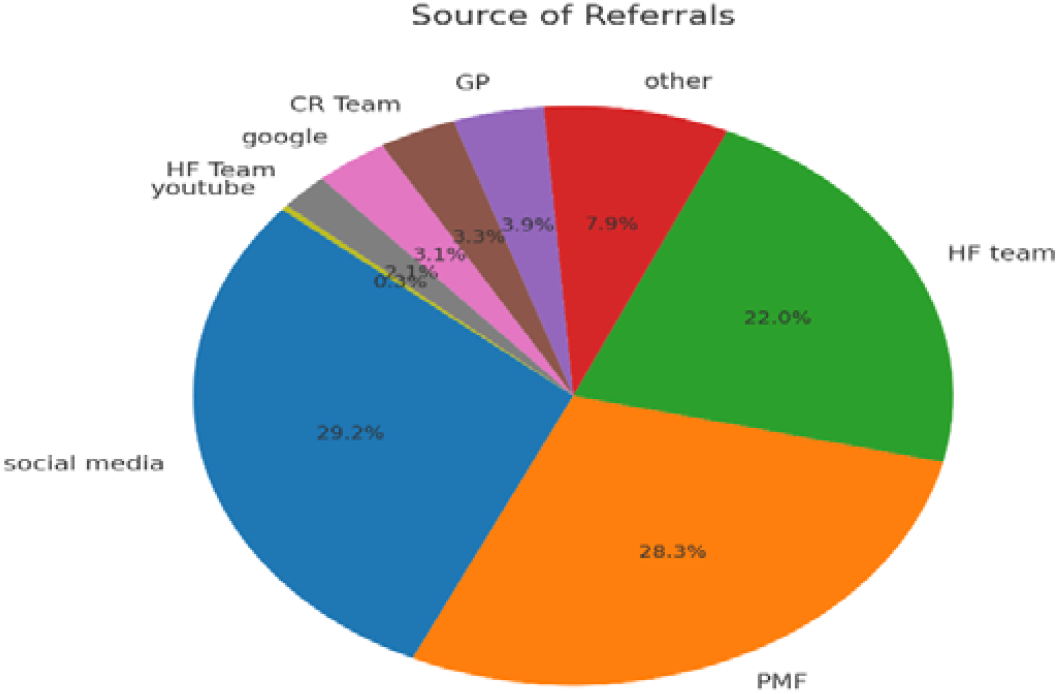
Sources of referrals to PMF CR portal.

Analysis of the time since HF diagnosis demonstrated late entry to CR: 343 (51%) registered >12 months post-diagnosis, 88 (13%) within 3 months, and 130 (19%) within 6–12 months. Only 38 (6%) reported any prior CR participation. We also correlated cardiac rehabilitation booklet orders from hospitals with National HF Audit HF admissions [9]. As shown in the scatter plot (Figure 2), within the limitations of the spread of the scatter, there was a general positive relationship — hospitals with more HF admissions tended to order more booklets. 33% of registrants came from the top 20 most deprived cities in England [10].

**Fig. 2.**
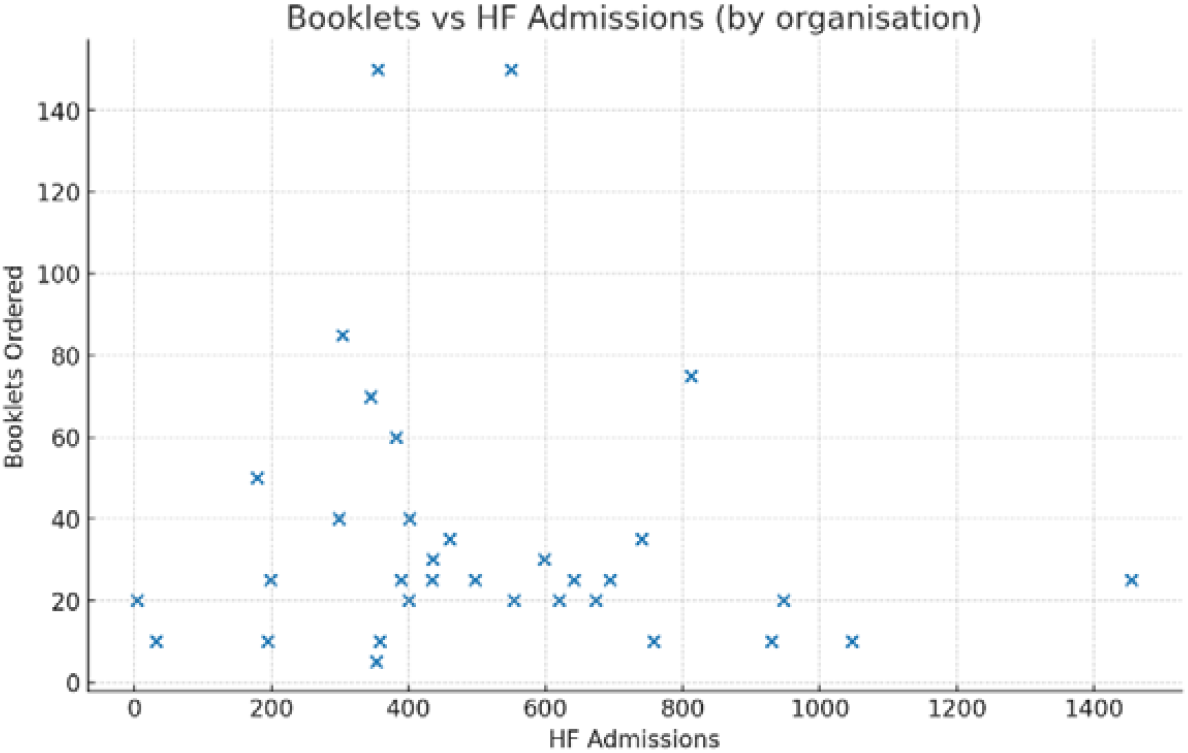
Scatter plot correlating number of orders for the PMF CR educational booklet with HF admissions.

## Discussion

This study provides crucial insights from the first year of roll-out of digital CR participation for HF using the PMF online platform. 94% of respondents had not participated in CR previously, and >50% joined 12 months after diagnosis. These findings highlight poor uptake of traditional CR and late engagement in standard CR, potentially due to difficulties in accessing hospital-based CR services, while also demonstrating the potential of digital platforms to institute early modification in physical activity, lifestyle and ensuring optimisation of therapies. This data also emphasises that PMF CR platform can play an essential role as an adjunct to standard cardiac rehab but also importantly as a life-long free offering, it can help bridge the gap in areas which may not have commissioning for standard cardiac rehab, providing a free service allows existing CR services to manage high acuity pts and alleviates resource as well as service burden. Digital platforms are a beneficial alternative for suitable patients by:

- benefiting people with learning styles different from the traditional hospital-based model
- allowing patients to progress through the rehabilitation programme at their own pacel
- allowing for ongoing lifelong revision of content not available through face-to-face rehabilitation programmes which have defined, time limited access.l

Our analysis also shows a gender pattern reversal, as, unlike traditional CR, where women are underrepresented [5,6], the majority of the online PMF CR cohort were women (63%). We can infer from this data that online, flexible formats and peer-led promotion may better suit women’s caregiving roles and accessibility needs. Socio-economic deprivation is linked with worse HF outcomes [11], and patients from these areas also have lower access to CR [5,6]. It was encouraging to note that up to a third of registrants who access the PMF CR portal were from the top 20 most deprived cities in England, indicating that access to this portal can help improve accessibility to CR in deprived parts of the country.

## Conclusions

By providing free lifetime access to online cardiac rehabilitation, widening the access of cardiac rehab to more women and people without access to standard cardiac rehabilitation (due to staffing, cost constraints, accessibility issues) and reaching areas with socio-economic deprivation, the PMF online cardiac rehab platform can help to increase CR uptake and reduce the inequity in access to CR in the UK.

## Data Availability

All data produced in the present study are available upon reasonable request to the authors

